# Clinical Evaluation of In House Produced 3D Printed Nasopharyngeal Swabs for COVID-19 Testing

**DOI:** 10.1101/2021.05.26.21257548

**Authors:** Simon Grandjean Lapierre, Stéphane Bedwani, François DeBlois, Audray Fortin, Natalia Zamorano Cuervo, Karim Zerouali, Elise Caron, Philippe Morency Potvin, Simon Gagnon, Nakome Nguissan, Pascale Arlotto, Isabelle Hardy, Catherine-Audrey Boutin, Cécile Tremblay, François Coutlée, Jacques de Guise, Nathalie Grandvaux

## Abstract

3D printed alternatives to standard flocked swabs were rapidly developed to provide a response to the unprecedented and sudden need for an exponentially growing amounts of diagnostic tools to fight the pandemics of COVID-19. In light of the anticipated shortage, an hospital-based 3D printing platform was implemented in our institution for the production of swabs for nasopharyngeal and oropharyngeal sampling based on the freely available open-sourced design made available to the community by University of South Florida’s Health Radiology and Northwell Health System teams as replacement for locally used commercial swabs. Validation of our 3D printed swabs was performed by a head-to-head diagnostic accuracy study of the 3D printed “Northwell model” with the cobas PCR Media swabs sample kit. We observed an excellent concordance (total agreement 96.8%, Kappa 0.936) in results obtained with the 3D printed and flocked swabs indicating that the in-house 3D printed swab can be used reliably in a context of shortage of flocked swabs. To our knowledge, this is the first study to report on autonomous hospital-based production and clinical validation of 3D printed swabs.

## Introduction

In December 2019, the outbreak of an atypical severe respiratory syndrome, now known as coronavirus 19 disease (COVID-19) in Wuhan, China (1), rapidly led to the identification of the Severe acute respiratory syndrome (SARS)-Coronavirus 2 (SARS-CoV-2). SARS-CoV-2 is highly transmissible (2, 3) and its rapid spread throughout the world led the World Health Organization to declare the COVID-19 outbreak a global pandemic on March 11^th^ 2020 (4). This triggered an unprecedented and sudden need for an exponentially growing amount of personal protection equipment and diagnostic tools. Real-time Polymerase Chain Reaction (PCR)-based detection of SARS-CoV-2 nucleic acids has rapidly become the gold standard to diagnose patients infected with SARS-CoV-2 (5). As typically performed for the detection of respiratory viruses, accurate COVID-19 PCR assay relies on the collection of samples from the upper respiratory tract including nasopharyngeal (NP) and oral mucosal surfaces (6). Flocked swabs feature perpendicular fibers that optimize specimen collection and elution in transport media and are hence considered optimal for sampling the respiratory tract mucosal surfaces. As testing rapidly became critical for the development of a COVID-19 response strategy, the world encountered shortage of PCR reagents and sampling swabs, resulting in testing backlogs, delayed diagnosis, compromised contact tracing and quarantine of patients and, potentially increased disease transmission.

The versatility of 3D-printing as well as the possibility of rapidly developing prototypes have enabled rapid mobilization of this technology to provide a response to the interruption of supply chains (7, 8). 3D printed alternatives to standard flocked swabs were rapidly developed. Early in the pandemic, open-sourced design for 3D printed swabs, were generously made available to the community by the teams from University of South Florida’s (USF) Health Radiology and Northwell Health System (9, 10). The design and workflow for hospital-based printing was subsequently published (8). Local manufacturing based on 3D printing is among the strategies that can help alleviate supply chain shortages.

In light of the anticipated shortage of sampling swabs in our hospital and in the province of Quebec, we sought to locally manufacture and evaluate sterile 3D printed swabs based on the freely available open source designs as replacement for locally used commercial swabs (8). Here, we report on the fabrication process and clinical evaluation of the 3D printed swabs against the cobas PCR Media□ swab sample kit in a prospective cohort of symptomatic healthcare workers. To our knowledge, this is the first study to report on autonomous hospital-based production and evaluation of 3D printed swabs. Our study showed that our locally printed swabs were a reliable alternative to commercial swabs and confirmed the initial assumption made by the USF and Northwell Health system groups that hospital-based production of 3D printed swabs can constitute a rapid, on-demand local response to provide replacements for pandemic-related disruption of the swabs supply chains.

## METHODS

### 3D printing

3D prototypes of swabs were designed and shared by teams from the Division of 3D Clinical Applications at USF and Northwell Health system (8). All final printed models successfully passed a complete set of mechanical tests (11). In our hospital-based production, we used the model referred to as “Northwell model” to which a breaking point located at 80 mm from the tip was added to the original design.

Swabs were printed by stereolithography (SLA) using Formlabs Form 3 and 3B printers (Formlabs, Somerville, Massachusetts, USA) with Surgical Guide resin (Formlabs, Cat#RS-F2-SGAM-01) that is biocompatible and sterilizable. The Preform software (Formlabs) was used to create an array of 256 swab models to be converted into printer instructions. The thickness of each printed layer was set to 0.1 mm. Printed swabs were cleaned in 99% isopropyl alcohol (IPA) (Sigma Aldrich, Cat#PX1835-3) for 20 minutes using the Form Wash (Formlabs) station and allowed to air dry for at least 30 minutes before being post-cured at 70°C for 30 minutes in a Form Cure (Formlabs) station. Visual inspection was performed to verify that printed swabs were free of defects, otherwise they were discarded.

### Sterilization

At the end of the production procedure, 3D printed swabs were immediately individually packed in autoclavable flat pouches (4 in × 10.5 in, Stevens Company, #164-S5) previously identified with autoclave-resistant laser labels (GA International, #AKA-13) indicating the swabs assigned lot number and date of production. Pouches were sealed with a vacuum rotosealer machine (Wipak Medical, #RS120). Appropriate sealing was verified visually. Sterilization was performed by autoclaving using a pre-vacuum steam cycle set at 132°C (270°F) for 4 min followed by a 30 min drying period.

To verify the efficacy of the sterilization procedure for each lot, the head of two swabs per production lot were inoculated with 10 μL of the Biological Indicator *Geobacillus stearothermophilus spore* suspension (1.7*10^7^ CFU/0.1 mL; Steris Corporation, #NA-091) under sterile conditions (12). Inoculated swabs were held horizontally for 30min and further let dry vertically for 24 h under sterile conditions. Next, dried swabs were sealed individually in sterilization pouches. One of the inoculated swabs was subjected to sterilization as described above, together with all swabs from the same lot. The other inoculated swab was kept in the pouch and was not sterilized. The two inoculated swabs were broken at the breakpoint and inserted into a bacteriology tube (Sarstedt, #62515006) containing 5 mL of tryptic soy broth culture media (Sigma Aldrich, #1463170010). Bacteria growth was monitored by incubation at 55°C with orbital agitation for 7 days. A culture media only negative control was also used. OD600nm was measured on day 7 to assess bacterial growth.

### Quality control

Swabs showing excessive warping post-sterilization were discarded after visual inspection. Basic mechanical testing was performed using a guide formed by 3 semicircular canals with a minimum radius of 15, 25 and 35 mm, respectively, that allowed testing the flexibility of swabs. The test was successful if the head and neck remained intact after going through each canal. The final test consisted in breaking the swab in half with one hand at its breaking point.

### 3D swabs clinical evaluation study participants’ recruitment

This study was performed in our institution’s COVID-19 rapid screening clinic and included symptomatic healthcare workers self-presenting for COVID-19 diagnostic testing. During an initial recruitment phase, participants were tested simultaneously with both the cobas PCR Media® swab sample kit (Roche Diagnostics, Laval, Quebec, Canada) and the 3D printed swabs. As for any COVID-19 test, all results were transmitted to the institution occupational health and safety office and public health authorities. In a subsequent recruitment phase, to increase the number of positive samples within the study, patients having previously tested positive on routine testing were contacted by clinical research personnel and offered to participate in the study. For those participants, both sampling techniques were repeated simultaneously.

### Oro-nasopharyngeal swab collection and SARS-CoV-2 PCR testing

A sequential oropharyngeal and nasopharyngeal sampling with each single swab was performed. For the nasopharyngeal sampling, Cobas PCR Media swabs sample kit and 3D printed swab were used in the same nostril in a randomized order. The swabs were transferred separately to tubes containing the cobas PCR Media transport medium before proceeding with the PCR analysis. All samples were tested on the FDA emergency use authorization (EUA) approved and locally validated cobas 8800 automated RT-PCR system which simultaneously tests the *ORF1 a/b* and *E*-gene viral molecular targets together with an internal control (13).

In conclusion, our study adds to the few clinical validation studies that demonstrated safety and accuracy of 3D swabs. Our study is unique in that it tested a fully integrated hospital-based production of 3D-printed swabs as initially suggested by the USF and Northwell Health system groups. Our clinical trial has demonstrated that our local 3D printed swab production line offers a reliable local alternative to commercial swabs and therefore confirms that it is a viable local response to provide replacements in the event of pandemic supply chain disruption.

### Statistical analysis

The Lilliefors statistical test adapted from the Kolmogorov–Smirnov nonparametric test was used to assess data distribution normality. The Wilcoxon matched-pairs test for non-normal distributions was used to evaluate the difference between means of RT-PCR cycle thresholds (Ct) obtained following both sampling methods. Concordance analysis between both assays using overall, positive and negative agreement percentages was performed with calculation of the Cohen’s Kappa values. By definition, Kappa values above 0.75 indicate excellent agreement, values between 0.40 and 0.75 indicate fair to good agreement, and values below 0.40 represent poor agreement beyond chance (14). Results obtained with the cobas PCR Media® swabs sample kit were considered as reference for positive and negative agreement calculation purposes.

### Ethical approval

The study received ethical approval from the Comité d’éthique de la Recherche of the Centre de Recherche du Centre Hospitalier de l’Université de Montréal (CHUM, 2021-9015 / 20.079). All participants were provided written informed consent.

## RESULTS

### 3D printed swab model

In a pilot print to start implementing a hospital production of 3D printed swabs, a biocompatible and sterilizable surgical guide resin to print two models from the designs shared by the 3D Clinical Applications Division of the USF and Northwell Health System was used. The two printed models are referred to as “USF” and “Northwell” (8). Based on the flexibility and the smaller head size, the “Northwell model” was selected by the clinical diagnostic team to pursue with the clinical tests. The cattail design of the “Northwell” model is composed of a head (18.0 mm long and 3.3 mm in diameter), a flexible neck (56.0 mm; 1.2 mm) and a handle (77.0 mm; 2.6 mm). Based on this pilot evaluation, a breaking point (1.0 mm; 1.4 mm) located at 80 mm from the tip to the original design was added to facilitate the release of the swab head in the transport tube containing the transport medium (**Figure 1A-C**). Three separate lots of 256 swabs were printed to ensure reproducibility of the production process and of the subsequent clinical evaluation. Swabs were packed individually in sterilization pouches before autoclaving (**Figure 1D**). Efficiency of the sterilization was verified for each lot through inhibition of *Geobacillus stearothermophilus spore* suspension inoculated on the head of one swab per lot (**Figure 2**). Two swabs from each printed lot were subjected to quality control check including testing of the flexibility (**Figure 1E**) and breaking of the swabs at the breaking point.

**Figure 1.**
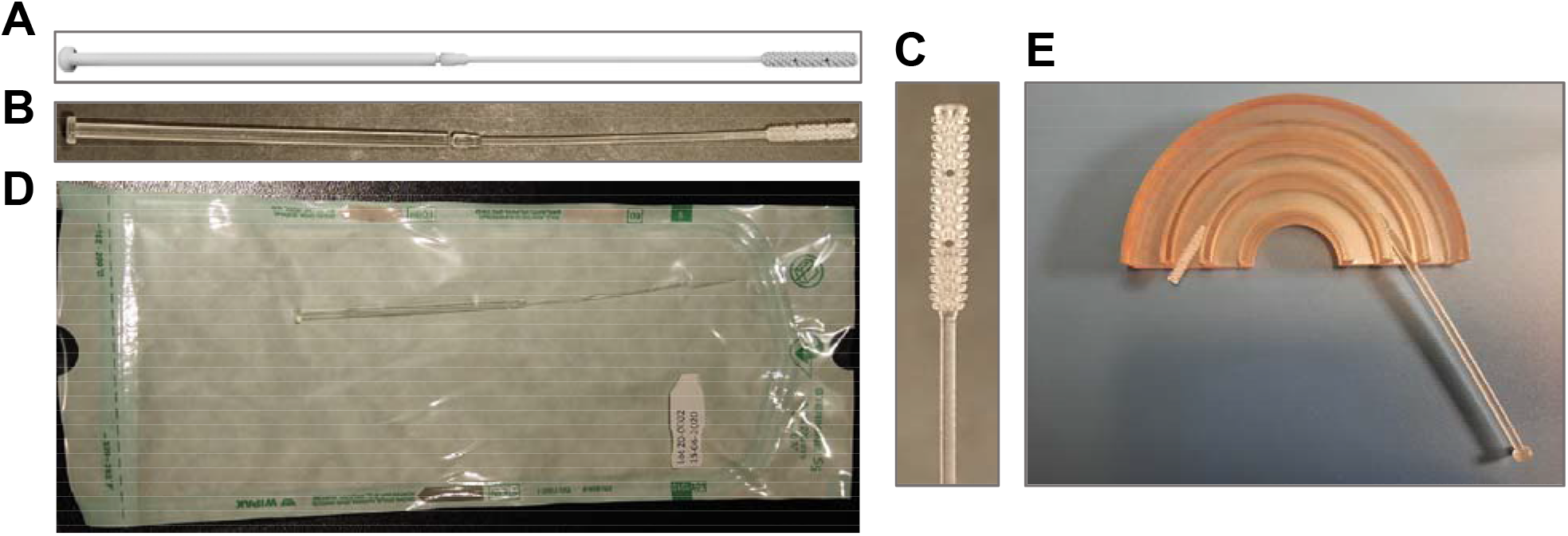
In-house 3D printed swab model. Design of the Northwell 3D swab model with the addition of a breakout point (**A**) used to 3D-print swab in our hospital (**B and C**). Swabs were individually packed in autoclavable and vacuum-sealed poches (**D**) for sterilization. (**E**) Flexibility was mechanically tested using semicircular canals (radius of 15, 25 and 35 mm).

**Figure 2.**
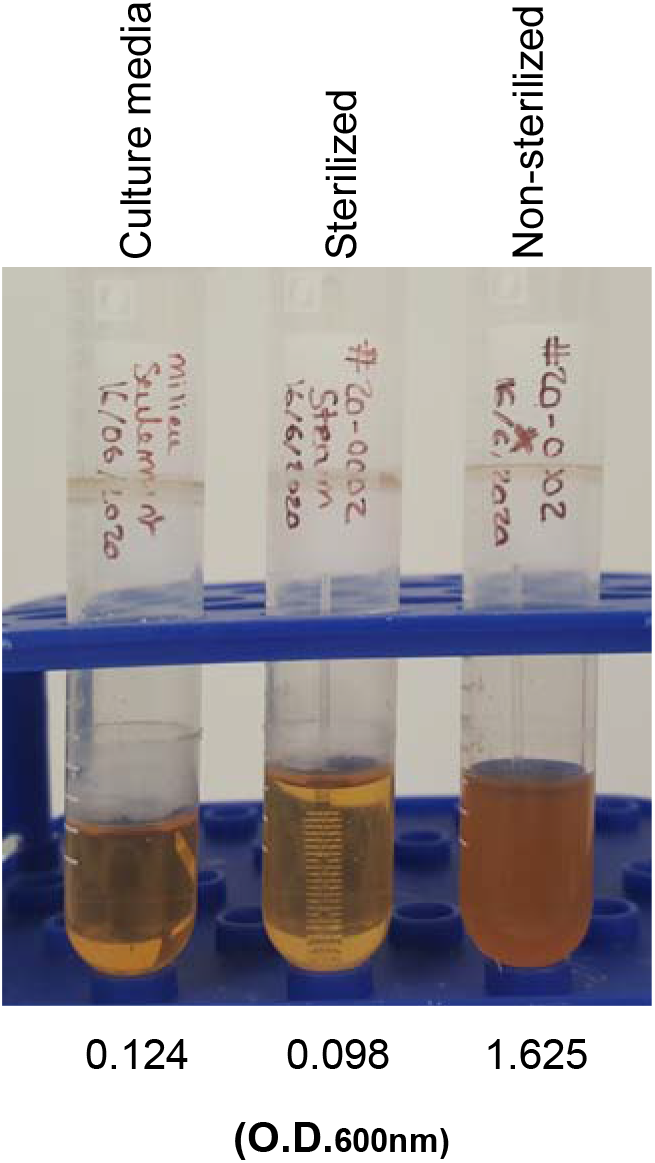
Validation of 3D printed swabs sterilization. Swab heads inoculated with *G. stearothermophilus* spore suspension before sterilization cultured in soy broth culture media. Bacteria growth was assessed by measuring the optical density (O.D.) at 600nm. Culture media alone was used as negative control.

### Clinical testing

A total of 63 participants were enrolled in the study. Thirty-two participants tested negative and 31 tested positive with the cobas 8800 SARS-CoV-2 PCR. Swabs from the 3 distinct printed lots were used for sample collection (batch 1, n = 21, 11 positives, 10 negatives; batch 2, n = 21, 11 positives; 10 negatives; batch 3, n = 21, 9 positives, 12 negatives). Lilliefors statistical test and Kolmogorov–Smirnov nonparametric test showed that cycle threshold (Ct) distributions for both swabs were not standard (p=0.12). PCR results for the *E-*gene (p=0.27), *ORF1* gene (p=0.92) and internal control (p=0.59) did not show significant Ct differences between flocked swabs and 3D printed swabs (**Figure 3 and Table 1**). A full agreement table between the flocked and the 3D printed swabs is presented in **Table 2A**. Overall, positive and negative agreements were respectively 96.8% (61/63), 96.9% (31/32) and 96.8% (30/31) with a Kappa value of 0.936 (**Table 2B**). In 2 cases, results obtained with the 2 swabs were discordant. In one case the flocked swab sample was positive with a Ct of 37.3 for the *E* gene only. For this same participant, the sample obtained with the 3D printed swab led to no amplification for both genes. In the second discordant case, the flocked swab sample was negative for both genes, while the 3D printed swab result was positive for both *ORF1* (Ct = 33.2) and *E* (Ct = 38.1) targets. Although, in both discordant cases the detected Ct were high, several other samples included in this study also had Ct in the high range without showing discordant results.

**Figure 3.**
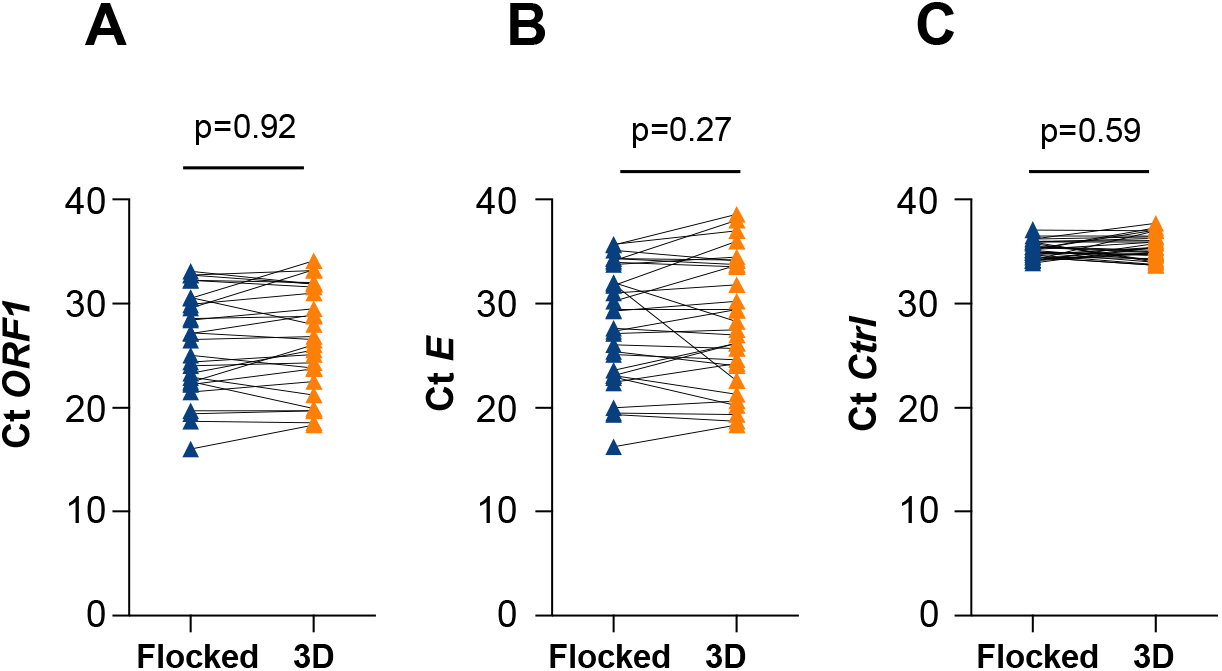
Cycle threshold (Ct) values of reverse-transcriptase polymerase chain reaction (RT-PCR) for the ORF1 and E genes. Participants were swabbed in the same nostril with a flocked and a 3D printed swab, successively. RT-PCR was performed to measure the Ct values of *ORF1* (A) and *E* (B) viral genes for each swab. An internal control (Ctrl) was also included (C). Statistical analyses are detailed in Table 1.

**TABLE 1.** Means Ct values of samples collected from individuals tested with a flocked or a 3D printed swab^a^.

|  | Flocked<br>mean Ct | 3D<br>mean Ct | Delta<br>Ct | p-<br>value |
| --- | --- | --- | --- | --- |
| ***ORF1*** | 26.06 | 26.51 | 0.44 | 0.92 |
| ***E*** | 28.03 | 28.30 | 0.26 | 0.27 |
| ***Ctrl*** | 35.16 | 35.35 | 0.19 | 0.59 |

<sup>a</sup>Difference between means was evaluated with the Wilcoxon matched-pairs test.

**TABLE 2.** Tables of agreement between flocked and 3D printed swabs showing positive and negative concordance percentages with Kappa values (**A**) and total number of positive (+), negative (-) and inconclusive (IC) PCR results (**B**)^b^.

| **A** |  |  |  |  |  |
| --- | --- | --- | --- | --- | --- |
|  |  | 3D | 3D | 3D | Total |
|  |  | + | - | IC |  |
| Flocked | + | 30 | 1 |  | 31 |
| Flocked | - | 1 | 31 | 0 | 32 |
| Flocked | IC | 0 | 0 | 0 | 0 |
| Total |  | 31 | 32 | 0 | 63 |

| **B** |  |  |
| --- | --- | --- |
|  | % | 95% IC |
| Positive concordance | 96,8 | 82,4 - 99,9 |
| Negative concordance | 96.9 | 82,9 - 99,9 |
| Total concordance | 96.8 | 88,5 - 99,8 |
| Kappa value | 0.936 | 0,738 - 0,994 |

<sup>b</sup> IC: Confidence Interval

## DISCUSSION

In this report, we described the implementation of a hospital-based 3D printing platform for the production of swabs for nasopharyngeal and oropharyngeal sampling for COVID-19 diagnosis. Such swabs are classified as Class I medical devices under the Canadian regulatory framework of Health Canada. To validate our 3D printed swabs, we performed a head-to-head diagnostic accuracy study of the 3D printed “Northwell model” swab (8) with the cobas PCR Media swabs sample kit. We observed a high concordance (total agreement 96.8%, Kappa 0.936) in results obtained with the 3D printed and flocked swabs indicating that the in-house 3D printed “Northwell model” swab can be used reliably in a context of shortage of flocked swabs. Previous clinical trial performed by the USF and Northwell Health System teams have reached similar conclusions by comparing the 3D printed “USF model” swab with standard flocked swabs using alternative transport medium, including the World Health Organization (WHO) approved viral transport media, media produced in-house according to the procedure described by the Centers for Disease Control and Prevention (CDC) or commercially available Universal Transport Media (10). At the time of starting our clinical trial, our hospital was not facing a shortage of supply, or planning to be out of stock in the medium term, of swabs for the diagnosis of COVID-19. Therefore, we were able to use the transport medium provided in the cobas PCR Media swabs sample kit with the 3D printed swab to ensure consistency in the clinical evaluation. The RT-PCR was performed through the trial on a single local cobas 8800 automated RT-PCR system authorized by Health Canada under an interim order for use related to COVID-19 diagnosis after complete published validation of the assay (13, 15). Given the complexity and discomfort associated with repeated simultaneous nasopharyngeal testing and the necessity to validate the use of 3D printed swab in a controlled head-to-head approach, we did not include other transport media in our validation study. We did not observe significant Ct values differences between swabs for the *ORF1* and *E* SARS-CoV-2 genes in the Roche cobas assay. These observations are in agreement with the data from the clinical trial performed by the USF and Northwell system teams using the 3D printed “USF model” swab compared to the Roche cobas sampling kit (97.03% agreement, Kappa 0.863) (10). In a study comparing swabs from four distinct 3D-printing manufacturers to the Copan swab (501CS01) using a RT-PCR run on a Abbott m2000 RealTime system platform, high degree of concordance with Kappa values between [0.85-0.89] were observed (16). An additional prospective clinical validation compared 3D-printed swabs from two manufacturers with the Universal Viral Transport Kit by Becton, Dickinson & Company using a liaison MDX RT-PCR machine (DiaSorin Molecular, LLC) and the Simplexa COVID-19 Direct Kit. Again, an excellent concordance between sampling procedures was observed (17).

To our knowledge, this is the first study reporting on autonomous hospital-based production and evaluation of 3D printed swabs. The implementation of our hospital-based platform was facilitated by the existing “Health-related 3D printing centre” in our institution, which normally serves the needs of the Radiation Oncology department and other hospital sectors, but quickly reoriented its efforts to provide healthcare workers with the much-needed protection equipment and diagnostic tools. Additionally, we had access to an institutional sterilization service that allowed us to easily perform the steam sterilization that was chosen as a rapid, nontoxic and inexpensive technique that is microbicidal and sporicidal, and was previously shown to be compatible with the Surgical Guide resin (18). Actual installations and staff availability allowed us to limit the production costs of 3D printed swabs. The total cost per swab was about 0.56USD, which included 0.26USD for consumables and 0.29USD for staff wages. Further optimization has yet to be realized to increase the production volume and lower the production costs.

In conclusion, our study adds to the few clinical validation studies that demonstrated safety and accuracy of 3D swabs. Our study is unique in that it tested a fully integrated hospital-based production of 3D-printed swabs as initially suggested by the USF and Northwell Health system groups. Our clinical trial has demonstrated that our local 3D printed swab production line offers a reliable local alternative to commercial swabs and therefore confirms that it is a viable local response to provide replacements in the event of pandemic supply chain disruption. In-house production of 3D printed swabs option remains particularly relevant as the need for testing capacity continues to increase across the world, as many countries are experiencing new waves of infection with the emergence of variants of SARS-CoV-2 (19, 20). Our experience in the rapid implementation of this production line could serve as an example for other institutions around the world in the fight against COVID-19.

## Data Availability

All data described in this manuscript will be provided upon reasonable request.

## ACKNOWLEDGEMENT

The authors thank Floriane Point and Stephanie Matte for help with the ethics approval and organization of patient recruitment. The authors thank all the patients who generously accepted to participate in this study and the laboratory technicians involved in this project. This study was funded by the TELUS Friendly Future Foundation/Fondation TELUS pour un futur meilleur and the Fondation du Centre Hospitalier de l’Université de Montréal. The funders had no role in the study design, data collection and interpretation, or the decision to submit the work for publication. SGL has received funding from Roche Diagnosis unrelated to the present study to test PCR Medium stability. FC received grants paid to the organization for research projects unrelated to the present study from Roche Diagnostics and Merck Sharp and Dome, honorariums for presentations from Merck Sharp and Dome and Roche diagnostics, and has participated in an expert vaccine group by Merck Sharp and Dome.

